# Joint effects of temperature and relative humidity on COPD hospital admissions in England, 2003-2021: A nationwide case-crossover study

**DOI:** 10.64898/2026.07.22.26358685

**Authors:** Di Xi, Arturo De la Cruz Libardi, Ana Catarina Pinho-Gomes, Bethan Davies, Antonio Gasparrini, Marta Blangiardo, Garyfallos Konstantinoudis

**Affiliations:** School of Public Health, Imperial College London; Environment & Health Modelling (EHM) Lab, Department of Public Health Environments and Society, London School of Hygiene and Tropical Medicine; Grantham Institute for Climate Change and the Environment, Imperial College London; The George Institute for Global Health, Imperial College London

## Abstract

**Background:** Heat exposure has been linked to chronic obstructive pulmonary disease (COPD) outcomes, but evidence on relative humidity and humidity-temperature interaction remains limited and inconsistent.

**Methods:** We conducted a nationwide time-stratified case-crossover study of 473,494 COPD hospital admissions in England during the summer months (June-August) from 2003 to 2021. Case-control pairs were linked to high-resolution daily maximum temperature and relative humidity data derived from HadUK-Grid and ERA5-Land, respectively. Exposure-response relationships were modelled using distributed lag non-linear models. Joint effects were examined using interaction models, including linear interaction, temperature spline by humidity category, and fully categorical specifications. Analyses were stratified by age group, sex, and socioeconomic deprivation.

**Results:** COPD admission risk increases steeply at higher temperatures following a J-shaped curve. The exposure-response for relative humidity is U-shaped with a relative risk of 1.08 (95% Confidence Intervals: 1.00 - 1.15) at the 99^th^ relative humidity percentile. We find limited evidence of effect modification by socioeconomic deprivation for both exposures. An estimated 1,040 (95%CI: 820 - 1,270) summer COPD admissions per year are attributable to non-optimal temperatures and 910 (95%CI: 680 - 1,160) to relative humidity. Higher relative risks are observed with concurrent high temperature and high humidity, but the evidence is weak.

**Conclusion:** High temperature and high relative humidity are associated with increased risk of COPD admission, with limited evidence of an interaction. Relative humidity should be considered alongside temperature in environmental risk assessment and targeted adaptation strategies, such as proactive heat-health advice for people living with COPD.

**Key Message:** *What is already known on this topic:*

- Higher ambient temperatures are consistently associated with an increased risk of COPD hospital admission, but little is known about the role of relative humidity.

*What this study adds:*

- Using 19 years of nationwide individual-level data on 473,494 COPD hospital admissions in England, we identified a J-shaped association with temperature and a U-shaped association with relative humidity. Around 1,000 summer COPD admissions per year were attributable to non-optimal temperature and around 900 to non-optimal relative humidity. Evidence of interaction was weak.

*How this study might affect research, practice and/or policy:*

- Relative humidity should be considered alongside temperature in environmental risk assessments, early warning systems, and heat-health action plans for people with COPD. Adaptation strategies, including housing improvements, indoor cooling, and tailored guidance, should be developed and evaluated to reduce temperature- and humidity-related COPD hospitalisations.

## Introduction

Chronic obstructive pulmonary disease (COPD) is the third leading cause of death worldwide and is characterised by recurrent exacerbations that frequently require hospitalisation^1^. In England, COPD imposes substantial burden on the National Health Service, accounting for over a hundred thousand emergency admissions each year, considerable healthcare costs, and a large number of years living with disability^2^. Acute exacerbations of COPD are multifactorial and are influenced by individual factors such as sex, age, disease severity, respiratory infections and comorbidities^3^. Environmental triggers, including air pollution and temperature, are increasingly recognised as important drivers of acute exacerbations^4,5^. However, evidence on the independent effects of relative humidity and its interaction with temperature remains limited.

Several epidemiological studies have reported increased risks of respiratory and COPD hospital admissions during periods of high temperature or heatwaves ^6–14^. Most previous work has treated temperature effects as linear or near-linear which does not fully characterise the complex exposure-response relationships. Evidence of the effect of relative humidity on hospital admissions for COPD is more limited and inconsistent, with studies variously reporting U-shaped, protective or monotonic associations, and only a few have examined the joint effects of temperature and humidity on COPD outcomes^9–16^. Moreover, ambient temperature and humidity can influence air pollution concentrations and chemistry, complicating the interpretation of their independent and joint effects^17,18^.

In this nationwide study in England restricted to summer months from 2003 to 2021, we investigated the association between daily maximum temperature, mean relative humidity and COPD hospital admissions using a time-series framework. We characterised the non-linear exposure-response relationships for temperature and humidity, examined their joint effects, and assessed effect modification by sex, age group and area-level socioeconomic deprivation. Finally, we explored the role of ambient air pollution, particularly PM_2.5_ and NO_2_, in shaping these associations.

## Methods

### Data

We analysed hospital admissions for COPD in England between 2003 and 2021 using Hospital Episode Statistics (HES) data retrieved from the Small Area Health Statistics Unit (SAHSU). For each admission, information on age group, sex, residential postcode at the time of hospitalisation, admission date, and discharge date was available. We included only admissions with a primary diagnosis code for COPD, defined by diagnostic codes J40-44 according to the International Classification of Disease V.10. (ICD-10)^19^. The analysis was restricted to summer months (June-August).

### Exposure

We used daily maximum temperature at a 1 km*×*1 km resolution from the UK Met Office with method described elsewhere^20^. We averaged the maximum temperature across lags 0–3 to capture cumulative health effects^21^.

Daily mean relative humidity was derived with established formulas from dewpoint and air temperature, both obtained from the ERA5-Land reanalysis dataset at a resolution of about 9km x 9 km ^22^. The rough daily mean humidity grid was resampled to match the resolution of the maximum temperature grid before exposure assignment.

### Covariates

We used particulate matter with a diameter of less than 2.5 µm (PM_2.5_) and nitrogen dioxide (NO_2_), in micrograms per cubic metre (µg/m^3^), as estimated by a daily 1 km x 1 km resolution hybrid ensemble machine-learning model^23^. To assess how deprivation modifies the effect of temperature and relative humidity, we retrieved the Index of Multiple Deprivation (IMD) at LSOA level in 2015 (Ministry of Housing, Communities and Local Government) and defined three categories of deprivation: most deprived (1^st^ quintile of IMD rank), intermediate (2^nd^, 3^rd^ and 4^th^ quintile of IMD rank) and least deprived (5^th^ quintile of IMD rank).

We further included wind speed and rainfall in the sensitivity analyses. We computed daily wind speed from the two wind component variables in ERA5-Land at a resolution of about 9 km x 9 km ^22^. Daily rainfall was obtained from Met Office at 1 km spatial resolution^20^. To assign daily exposures to health records, residential postcode centroids of each patient were spatially linked to the 1 km*×*1 km grid cell, applying a 100 m fuzziness to the postcode location to fulfil governance requirements.

### Statistical analysis

#### Study design

We used a time-stratified case-crossover design to study the transient exposure effects. For each COPD admission (the event), we compared the temperature on the event day (defined by admission dates) with temperatures on matched non-event (control) days for the same person. Because each person acts as their own control, this approach automatically adjusts for factors that don’t change or change slowly over time (for example, sex or socioeconomic deprivation). We sampled control days matching the day of the week and calendar month and year as the event day. This accounts for long-term trends, seasonality and overlap bias. Discharge dates ensured that control days were selected only from periods when individuals were not in hospital, to minimise exposure misclassification.

#### Model specifications

To allow for flexible effects of temperature and relative humidity, we used natural cubic splines with 3 knots on the 10^th^, 50^th^ and 90^th^ exposure percentile^24^. We reported both unadjusted models (each exposure included on its own) and fully adjusted models (including both exposures plus national holidays). In both unadjusted and fully adjusted models, we used a cluster robust variance-covariance matrix, clustering by patient, to account for recurrent hospital admissions. We fitted the models stratified by age (0-64 years, 65-74 years, 75 years and older) and sex and explored the interactions with deprivation. We also explored the role of air pollution by adding NO_2_ and PM_2.5_ (lag 0-3) separately in the fully adjusted models.

To study how temperature and relative humidity interact, we fitted three models. In model 1 (linear interaction) we included non-linear terms for temperature and relative humidity, as well as a linear interaction term between them. In model 2 we grouped relative humidity into three categories defined using percentiles low (lower than the 50% percent); medium (50-75%); and high (higher than 75%) and allowed non-linear temperature effects within each category. We modelled non-linear effects in model 1 and 2 using the same spline setup as described above. Lastly, in model 3, we split temperature and relative humidity into nine categories (using the percentiles defined earlier, i.e., low temperature/low humidity, low temperature/mid humidity, low temperature/high humidity, etc.) and estimated the relative risks on these categories using the low temperature/low humidity group as the reference.

Results were reported as means and 95% Confidence Intervals (CI) of the relative hospitalisation risk compared with the minimum hospitalisation threshold. The threshold was defined, for each exposure, as the value of maximum temperature or relative humidity within the 25^th^ to 90^th^ percentile range at which the relative risk is lowest^25^. This approach avoided unstable estimates at the extreme tails and allows identification of a more epidemiologically plausible minimum-risk point. For the linear interaction term, we reported mean and 95%CI of the % increase in the risk of COPD hospital admissions.

### Sensitivity analysis

In sensitivity analyses, we assessed the robustness of the exposure-response curves by additionally including rainfall and wind speed as linear terms, and by extending the lag period to 0-5 days, both in the fully adjusted model.

## Results

### Outcome

We identified 2,252,903 hospital admission records with COPD as the primary diagnosis between 2003 and 2021, Figure 1. After excluding 8,037 records of admissions to English hospitals occurring in people living in Wales, 1,769,807 records outside the summer months, 1,507 records with missing information and 58 duplicated records or records for which controls could not be sampled because the individuals were not discharged during the month, there were 473,494 records remaining available for the analysis.

**Fig 1.**
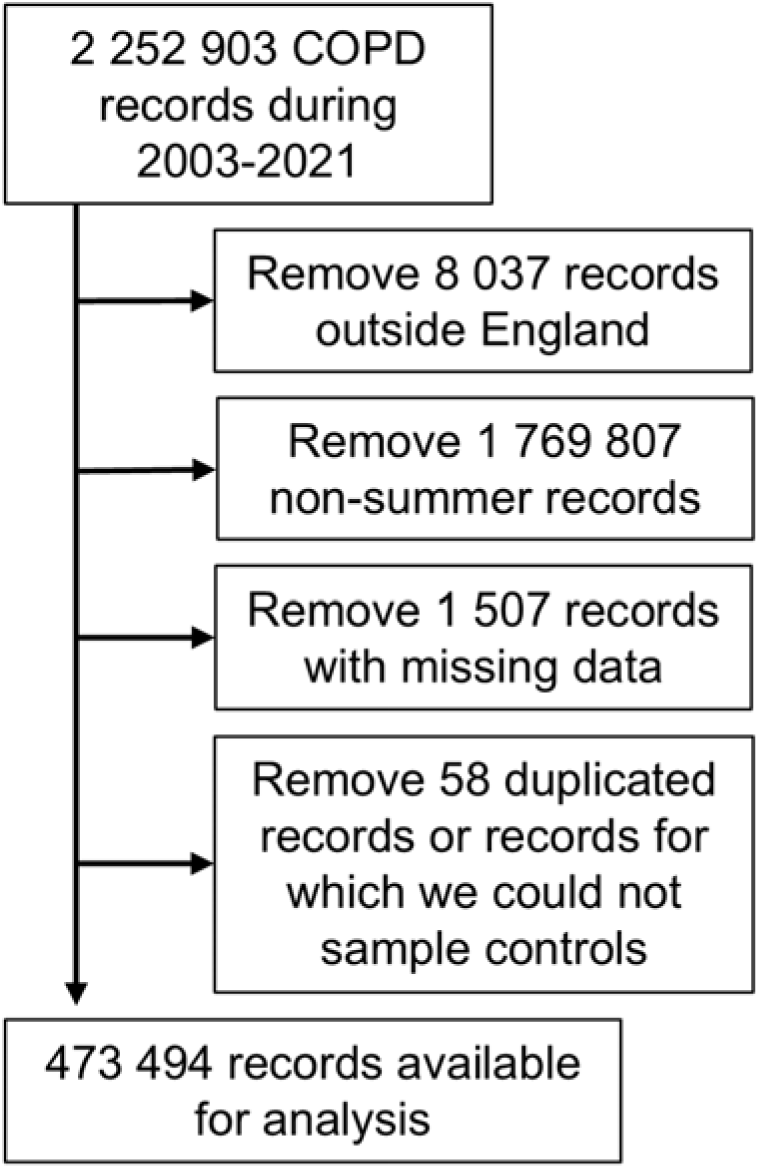
Flowchart of COPD hospital admissions selection criteria.

### Covariates

Online Supplement Table S1 shows the mean and standard deviation of maximum temperature, relative humidity, PM_2.5_ and NO_2_ by year and case-control status. The mean of the maximum temperature across England was 22.33^°^C among the controls and 22.57^°^C among cases in 2003. These values were slightly lower in 2021 at 21.15^°^C and 21.17^°^C respectively. The mean relative humidity across England was approximately 73% in 2003 and 77% in 2021 for both cases and controls. A downward trend in PM_2.5_ and NO_2_ concentrations is observed across the study period (Online Supplement Table S1).

### Overall results

Panel A of Figure 2 shows unadjusted and fully adjusted exposure-response associations for maximum temperature and relative humidity. For temperature, the minimum-risk threshold (MRT), defined as the temperature associated with the lowest hospitalisation risk, is approximately 20^°^C. There is no evidence of increased COPD admission risk below the MRT; above the MRT, risk rises steeply with an approximately exponential increase. These patterns are robust in the fully adjusted model. In the fully adjusted models and across all age and sex groups, the relative risks at the 90^th^, 95^th^ and 99^th^ temperature percentiles, each compared with the MRT, are 1.13 (95% Confidence Intervals (CI): 1.11-1.15), 1.22 (95% CI: 1.19-1.25) and 1.42 (95% CI: 1.34-1.52) respectively (Online Supplementary Table S2). The exposure-response for relative humidity is U-shaped. The minimum-risk relative humidity (MRH), defined as the relative humidity associated with lowest hospitalisation risk, is approximately 60%. The elevated COPD admission risk at both low and high humidity and greater uncertainty at the extremes. In the fully adjusted models and across all age and sex groups, relative risks at the 90^th^, 95^th^ and 99^th^ percentiles, compared with the MRH, are 1.07 (95% CI: 1.05-1.09), 1.07 (95% CI: 1.04-1.10) and 1.08 (95% CI: 1.00-1.15) respectively (Online Supplementary Table S2).

**Fig 2.**
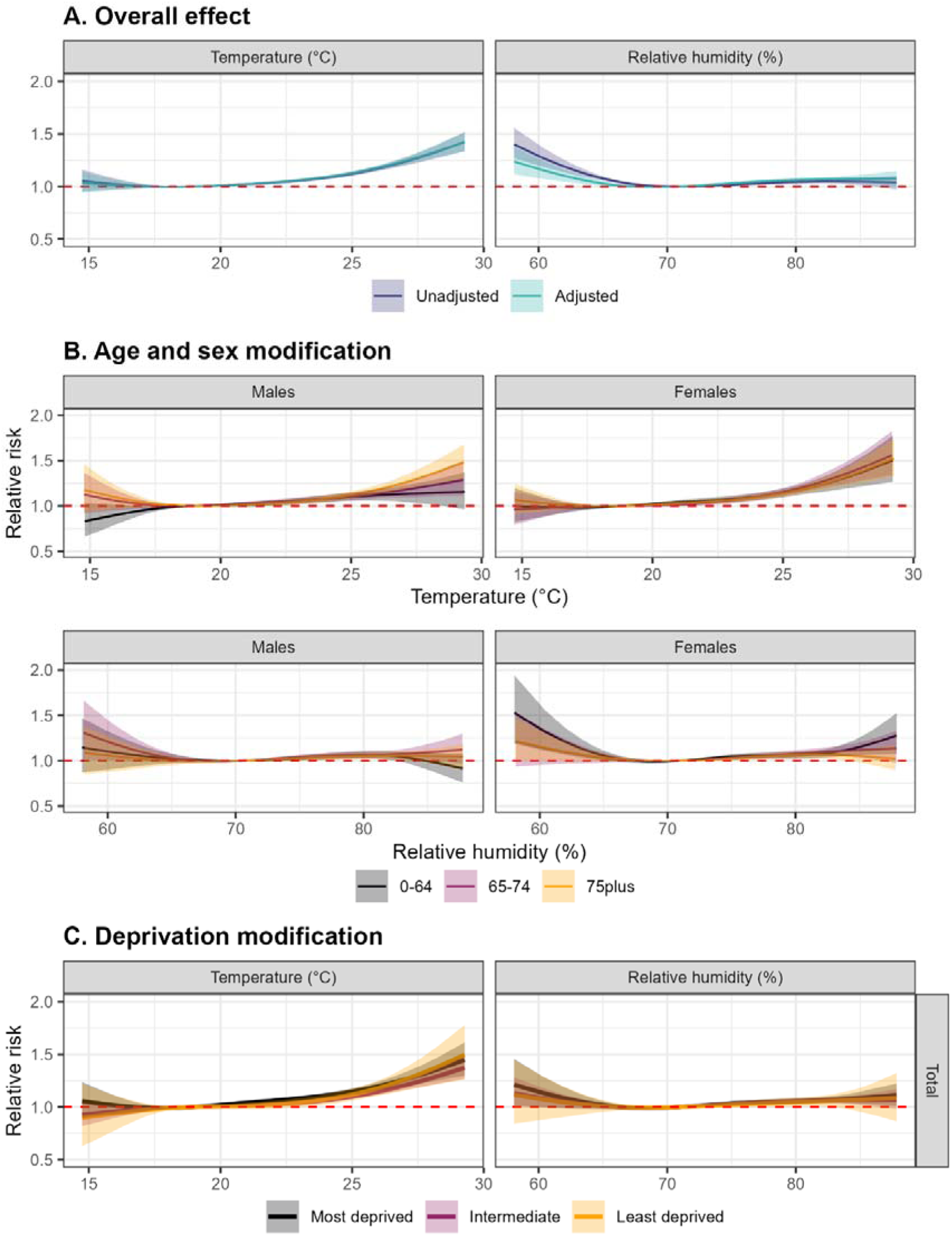
Exposure-response associations between maximum temperature, relative humidity, and COPD hospital admissions in England (2003–2021) *Footnote: Panel A represents the unadjusted and fully adjusted exposure-response curves for maximum temperature and relative humidity. Panel B represents the fully adjusted exposure-response associations stratified by age group and sex, while panel C represents the fully adjusted exposure-response associations across area-level socioeconomic deprivation categories*.

### Age and sex modification

Panel B of Figure 2 shows fully adjusted associations for maximum temperature and mean daily relative humidity stratified by age and sex. For temperature, women experience higher heat-related risk than men: at the 99^th^ temperature percentile, the relative risk is 1.54 (95% CI: 1.40-1.67) for women and 1.32 (95% CI: 1.21-1.45) for men in the total age group (Online Supplementary Table S2). Among men, heat-related risk increases with age, whereas a comparable age gradient is not evident among women. For relative humidity, we also observe sex-specific modification: at the 99^th^ percentile (all ages combined), men have lower risk (1.03; 95% CI: 0.95-1.12) than women (1.12; 95% CI: 1.03-1.22) in the total age group. Among women, the highest humidity-related risks are observed in the youngest age group, while in men there is no clear evidence of age modification (Panel B Figure 2).

### Deprivation modification

Panel C of Figure 2 shows the non-linear effects of COPD hospital admission risk in relation to maximum temperature and relative humidity across different deprivation categories. Overall, the analysis shows no evidence that the effect is modified by deprivation level.

### Health impact assessment

Table 1 shows the number of yearly COPD hospital admissions (rounded to the closest 0 or 5) attributable to non-optimal summer temperature and non-optimal summer relative humidity, stratified by age group and sex. Overall, an estimated 1,040 (95% CI: 820-1,270) summer COPD admissions per year can be attributable to non-optimal temperature and 910 (95% CI: 680-1,160) to relative humidity exposure. There were 460 (95% CI: 300-620) COPD hospitalisations attributable to non-optimal temperature and 380 (95% CI: 190-580) COPD hospitalisations attributable to non-optimal relative humidity per year among males, compared with 580 (95% CI: 430-730) and 530 (95% CI: 360-690), respectively, per year among females. (Table 1)

**Table 1.** Number of COPD hospitalisations attributable to non-optimal temperature and relative humidity in England from 2003 to 2021 (rounded to one significant figure)

| Sex | Age group | Total number of hospitalisations | Yearly hospitalisations attributable to temperature | Yearly hospitalisations attributable to relative humidity |
| --- | --- | --- | --- | --- |
| Males | 0-64 | 58,990 | 60 (-30, 140) | 80 (-10, 170) |
|  | 65-74 | 76,410 | 170 (80, 250) | 170 (60, 270) |
|  | ≥75 | 98,860 | 240 (120, 340) | 140 (0, 270) |
|  | Total | 234,260 | 460 (300, 620) | 380 (190, 580) |
| Females | 0-64 | 61,460 | 150 (80, 230) | 170 (90, 250) |
|  | 65-74 | 73,120 | 160 (70, 240) | 170 (70, 270) |
|  | ≥75 | 104,660 | 270 (160, 360) | 190 (70, 300) |
|  | Total | 239,230 | 580 (430, 730) | 530 (360, 690) |
| Total | 0-64 | 120,450 | 210 (100, 320) | 260 (130, 360) |
|  | 65-74 | 149,530 | 330 (200, 450) | 340 (200, 470) |
|  | ≥75 | 203,510 | 500 (350, 640) | 320 (160, 500) |
|  | Total | 473,490 | 1,040 (820, 1,270) | 910 (680, 1,160) |

### Synergistic effects

Figure 3 shows the joint effects of maximum temperature and relative humidity using the three model specifications described in the Methods. In Panel A, a linear interaction was fitted; evidence of interaction is weak overall, although estimates suggest higher joint risks in men. Panel B shows the non-linear temperature effects across relative humidity quantiles. Uncertainty is substantial and the overall evidence remains limited, but in men heat-related risk tends to increase at higher humidity quantiles. Panel C presents ratios of relative risks for each temperature-humidity category, using low temperature/low humidity as the reference. A general increase in risk is observed with higher temperature, particularly at medium to high humidity levels. For example, at low humidity, relative risks increase from 1.01 (95% CI: 0.99-1.02) at medium temperature to 1.06 (95% CI: 1.05-1.08) at high temperature, while at medium humidity, risks rise from 1.03 (95% CI: 1.02-1.05) to 1.08 (95% CI: 1.06-1.10). A similar pattern is also observed with higher humidity, with relative risks of 1.03 (95% CI: 1.02-1.04), 1.04 (95% CI: 1.02-1.06), and 1.07 (95% CI: 1.04-1.09) across low, medium, and high temperature categories, respectively. However, differences between categories are small and associated with overlapping confidence intervals, indicating uncertainties (Online Supplementary Table S3).

**Fig 3.**
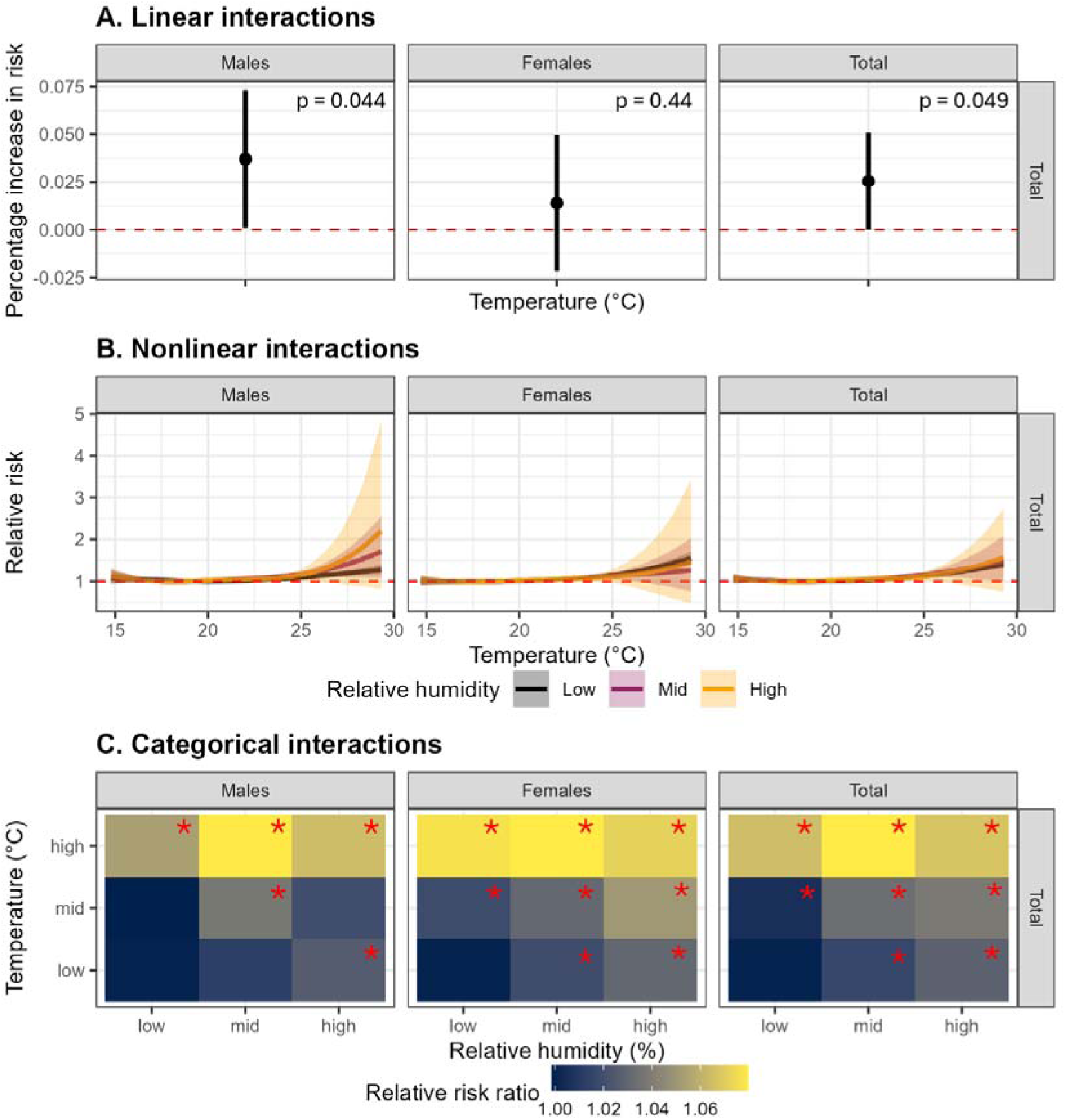
Joint effects of maximum temperature and relative humidity on COPD hospital admissions in England (2003–2021) *Footnote: Panel A represents the linear interaction model between temperature and relative humidity, with p-values shown for the interaction term. Panel B represents the non-linear temperature effects across relative humidity quantiles. Panel C represents the relative risks for combinations of temperature and humidity categories, with low temperature/low humidity as the reference; asterisks indicate statistically significant estimates with p-values < 0.05*.

### Air pollution adjustment

Adjustment for air pollution attenuates the temperature-admission association, with the greatest attenuation observed after controlling for PM_2.5_ (Figure 4). In contrast, the humidity-admission association is not affected by air pollution adjustment as the relative humidity exposure-response curves remain broadly unchanged (Figure 4).

**Fig 4.**
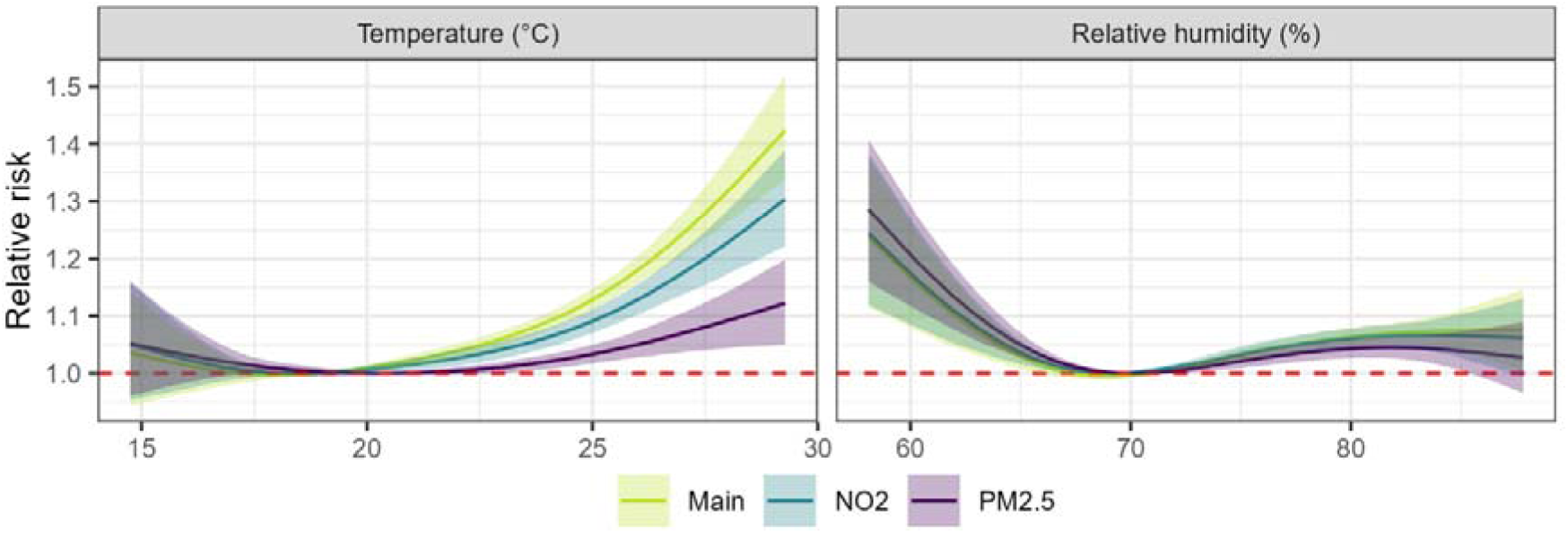
Sensitivity of temperature– and humidity–COPD hospital admission associations to adjustment for ambient air pollution in England (2003–2021)

### Sensitivity analysis

The exposure-response curves for temperature and relative humidity remain stable after adjustment for rainfall and wind speed (Online Supplement Figure S1). Extending the lag period to 0–5 days results in a modest amplification of risk at the extremes of maximum temperature, whereas the estimates for relative humidity are essentially unchanged (Online Supplement Figure S2).

## Discussion

In this nationwide study in England restricted to summer months, we find a J-shaped association between daily maximum temperature and COPD hospital admissions, with minimal risk at 20^°^C and a steep increase at higher temperatures. Heat-related risks are greater in women than men, and in men increase with age. Relative humidity shows a U-shaped association of smaller magnitude and women appear more vulnerable than men, with little evidence of age modification among men. We also find no evidence of an effect modification by deprivation for either exposure. Joint analyses provide weak overall evidence but suggest higher risks when higher temperatures coincided with higher relative humidity. Adjustment for ambient air pollution, particularly PM_2.5,_ attenuates heat-related risks, while associations with relative humidity are largely unchanged.

Rising summer temperatures have consistently been linked to higher risks of respiratory hospital admissions^26^. In Greater London, a 2000-2009 study reported an exposure-response shape between mean temperature and admissions for lower respiratory diseases similar to our findings^8^. Studies specifically examining COPD hospitalisations have most often modelled temperature as a linear or piecewise linear exposure. In Brazil, one such study found that during the four hottest months, the odds of COPD hospitalisation increased by 5% (95% CI 4% to 6%) for every 5°C increase in the average temperature (0-3 day lags), with women and older adults being the most vulnerable groups^6^. Another study, using data that partially overlap with ours, estimated a 1.5% increase in COPD hospital admission risk per 1°C increase in the maximum temperature (0-2 day lags), and attributed more than 1,800 events per year in England to temperatures above 23°C^7^. However, our analyses estimated approximately 1,000 COPD hospitalisations per year in England attributable to non-optimal summer temperatures. This discrepancy is plausibly explained by methodological differences, including the lag structure, study period, temporal changes (e.g., in clinical coding practices and population vulnerability), and the functional form used to characterise the exposure-response relationship.

Experimental studies suggest that humidity may affect respiratory function by decreasing lung capacity and increasing airway resistance^27,28^, but epidemiological evidence on relative humidity and health risks is mixed. A multi-country study spanning more than 20 countries reported weak evidence of a protective association between higher humidity and summer mortality after adjusting for temperature^15^, and city-level analyses in China similarly found decreasing mortality with increasing humidity^11^. Far fewer studies have evaluated hospital admissions. In Macau, China, total admissions rose sharply when relative humidity exceeded 90%^10^. Our COPD-specific findings are in keeping with studies reporting U-shaped associations for admissions, including work from Taipei^9^, but contrast with reports from Poland suggesting a protective effect^14^ and with studies in mainland China indicating a monotonic decrease in risk with higher humidity^16^. Differences in climate, population susceptibility, exposure assessment, confounder adjustment, lag structures, and model specification likely contribute to this heterogeneity across settings.

Fewer studies have investigated the joint effects of temperature and humidity on health. Some have approached this by using composite indices that combine these two exposures. For example, a study in Hefei, China, examining the impact of humidex on childhood asthma hospital admissions reported a U-shaped association^12^. Similarly, a study of twelve European cities found that respiratory hospital admissions increased with higher maximum apparent temperatures^13^. Our work is more closely aligned with studies that explicitly model the interaction between temperature and relative humidity, although findings from this literature are mixed. A large study across 353 locations in China (2006–2017) reported that dry-hot events were associated with a higher mortality risk than wet-hot events^11^. A multi-country analysis suggested that the mortality risk associated with high temperatures may be weaker under higher relative humidity^15^. By contrast, a Chinese study of ambulance dispatches reported that higher humidity amplified temperature-related health risks^29^. This conflicting evidence may be explained by differences in the population and percentage of humidity. Acclimatisation and environmental factors (e.g., availability of air conditioning and green and blue space) likely reduce the health impact of temperature but especially humidity. In line with pre-existing literature, we find higher temperatures increased the risk of COPD hospitalisation particularly for women and older individuals. Men appear to have an increased risk of hospitalisation on humid days, whereas the evidence for women is insufficient. The association between temperature and age is well established and likely reflects not only reduced physiological reserve and thermoregulation but also reduced ability to implement adaptation behaviours in response to heat and humidity. On the other hand, sex differences in heat- and humidity-related hospital admission, particularly the underlying mechanisms in females, is not well understood. These are likely to arise from a combination of anatomical, physiological (such as sex-specific gene regulation and inflammatory responses^30^) and social factors, including differences in smoking patterns and occupational or household exposures^31^.

We found no evidence of a deprivation gradient in temperature- or relative humidity-related COPD hospitalisation risk. Our finding is consistent with a previous nationwide case-crossover study in England, which also found little evidence that IMD modified heat-related COPD risk^7^. Evidence on socioeconomic status as a modifier of humidity-related COPD risk is even more limited. Our finding that deprivation did not modify the association with relative humidity adds to a sparse literature and should be interpreted cautiously. They may reflect the social patterning of COPD, as the condition is more common in socioeconomically disadvantaged groups^32^, which may reduce variation across deprivation levels within this population. Universal access to NHS care may also reduce socioeconomic differences in COPD management and vulnerability to exacerbations. In addition, IMD is an area-level proxy for individual deprivation. The stratified analysis by IMD level may not fully capture individual differences in housing conditions or adaptive capacity within the IMD strata.

However, the absence of effect modification by IMD for temperature or relative humidity should not be interpreted as indicating that socioeconomic deprivation has no role in COPD risk. Given the high underlying risk in people with COPD, socioeconomic disadvantage may still contribute to a greater burden of acute care use in this population, as shown by recent UK evidence on individual-level material deprivation and emergency healthcare utilisation^33^.

Ambient air pollution is a well-established determinant of COPD hospitalisation risk^34^. In our primary analyses, we treated air pollution as a potential mediator of heat-related effects and therefore did not adjust for it. In a subsequent analysis where we included PM_2.5_ and NO_2_ in the model, the estimated non-linear temperature association was attenuated, with the largest reduction observed after controlling for PM_2.5_. This pattern aligns with a previous study reporting attenuation of heat effects following pollution adjustment^35^.

A key strength of this study is the granular spatial linkage between outcomes and exposures. Using full residential postcodes, we connected COPD admissions to meteorological and air-quality data at high geographic resolution, reducing exposure misclassification from spatial misalignment. We ascertained hospital admissions from NHS England, which offers near-complete coverage of public sector hospitalisations in England, yielding a large, nationally representative dataset spanning 2003-2021 and thereby minimising selection bias related to differential healthcare access.

This study has several limitations. Assigning outdoor environmental conditions at the residential postcode is an indirect proxy for personal exposure; individuals move across multiple microenvironments and indoor conditions often diverge from ambient, so nondifferential exposure misclassification is likely and may attenuate associations. Despite adjustment for major environmental determinants of COPD hospitalisations, unmeasured and time-varying factors, such as aeroallergens, circulating respiratory infections, behavioural adaptation, and housing characteristics, could not be incorporated, leaving the possibility of residual confounding.

## Conclusion

In this nationwide study of summer COPD hospitalisations in England, we report that nonoptimal temperature and humidity impose a substantial burden, with clear potential to worsen under climate change. Women and older adults were consistently the most vulnerable groups. These findings highlight the need for targeted adaptation strategies for people living with COPD, including personalised heat-health advice, and improvements in housing and indoor environments. Proactive mitigation of heat and humidity exposures in the high-risk groups, such as targeted weather-health alert, should be a priority for public health and respiratory care planning.

## Data Availability

This work uses data provided by patients and collected by the NHS as part of their care and support. Hospital Episode Statistics data are copyright 2026, re-used with the permission of NHS England. All rights reserved.
The work of the UK Small Area Health Statistics Unit is overseen by UK Health Security Agency (UK HSA). This work was supported by Health Data Research UK, an initiative funded by UK Research and Innovation, Department of Health and Social Care (England) and the devolved administrations, and leading medical research charities. This paper does not necessarily reflect the views of UKHSA, NIHR or the Department of Health and Social Care. SAHSU does not have permission to supply data to third parties.

## Contributors

G.K. conceived and supervised the study, including model development, data acquisition and quality control. D.X. developed code for the statistical analyses and drafted the manuscript. A.D.C.L. processed and curated the exposure data. A.C.P.-G., B.D., A.G. and M.B. contributed to interpretation of the results and critical revision of the manuscript. All authors reviewed and approved the final version.

## Data sharing

SAHSU does not have permission to supply data to third parties.

## Acknowledgements

G.K. is supported by an Imperial College Research Fellowship. A.G. is supported by the Wellcome Trust (Grant IDs: 320878/Z/24/Z and 308914/Z/23/Z).

All authors acknowledge Infrastructure support for the Department of Epidemiology and Biostatistics provided by the NIHR Imperial Biomedical Research Centre (BRC NIHR203323).

This work uses data provided by patients and collected by the NHS as part of their care and support. Hospital Episode Statistics data are copyright © 2026, re-used with the permission of NHS England. All rights reserved.

The work of the UK Small Area Health Statistics Unit is overseen by UK Health Security Agency (UK HSA). This work was supported by Health Data Research UK, an initiative funded by UK Research and Innovation, Department of Health and Social Care (England) and the devolved administrations, and leading medical research charities. This paper does not necessarily reflect the views of UKHSA, NIHR or the Department of Health and Social Care.

## Ethics

The analyses were covered by the national research ethics approval from the London-South East Research Ethics Committee (Reference 22/LO/0256). Data access was covered by the Health Research Authority Confidentiality Advisory Group under section 251 of the National Health Service Act 2006 and the Health Service (Control of Patient Information) Regulations 2002 (Reference 20/CAG/0028).

