## Supplementary Materials for "Joint effects of temperature and relative humidity on COPD hospital admissions in England, 2003-2021: A nationwide case-crossover study"

**Affiliations**

**Table S1.** Mean and standard deviation (SD) of maximum temperature, relative humidity, PM_2.5_ and NO_2_ concentration by year and case-control status.

| Year | Status | Temperature | | Relative Humidity | | PM_2.5_ | | NO_2_ | |
| --- | --- | --- | --- | --- | --- | --- | --- | --- | --- |
|  |  | Mean | SD | Mean | SD | Mean | SD | Mean | SD |
| 2003 | Control | 22.33 | 3.06 | 73.13 | 6.25 | 12.46 | 4.69 | 22.17 | 8.72 |
| 2003 | Case | 22.57 | 3.25 | 73.06 | 6.30 | 12.99 | 5.22 | 22.54 | 8.95 |
| 2004 | Control | 20.89 | 2.77 | 76.00 | 5.16 | 9.82 | 2.86 | 21.06 | 7.82 |
| 2004 | Case | 20.97 | 2.81 | 76.13 | 5.27 | 9.95 | 2.98 | 21.18 | 7.89 |
| 2005 | Control | 20.89 | 2.98 | 74.77 | 5.11 | 8.13 | 2.34 | 19.11 | 8.38 |
| 2005 | Case | 20.95 | 3.06 | 74.77 | 5.16 | 8.19 | 2.39 | 19.15 | 8.49 |
| 2006 | Control | 22.47 | 3.64 | 71.73 | 6.29 | 9.66 | 3.45 | 20.52 | 9.98 |
| 2006 | Case | 22.69 | 3.72 | 71.67 | 6.43 | 9.89 | 3.62 | 20.90 | 10.05 |
| 2007 | Control | 19.73 | 2.00 | 78.14 | 5.45 | 7.02 | 2.37 | 18.90 | 8.33 |
| 2007 | Case | 19.82 | 2.01 | 78.34 | 5.34 | 7.13 | 2.46 | 19.04 | 8.35 |
| 2008 | Control | 20.12 | 2.29 | 76.04 | 5.13 | 9.26 | 2.45 | 18.04 | 7.85 |
| 2008 | Case | 20.11 | 2.31 | 76.1 | 5.09 | 9.31 | 2.49 | 18.11 | 7.88 |
| 2009 | Control | 20.81 | 2.71 | 74.76 | 5.12 | 9.90 | 3.35 | 17.85 | 8.41 |
| 2009 | Case | 20.86 | 2.78 | 74.71 | 5.10 | 10.01 | 3.39 | 18.03 | 8.51 |
| 2010 | Control | 20.85 | 2.65 | 73.17 | 6.79 | 9.82 | 2.97 | 18.20 | 7.57 |
| 2010 | Case | 20.85 | 2.66 | 73.22 | 6.80 | 9.94 | 3.12 | 18.35 | 7.68 |
| 2011 | Control | 19.83 | 2.24 | 73.57 | 5.33 | 9.43 | 2.27 | 18.70 | 7.85 |
| 2011 | Case | 19.88 | 2.28 | 73.64 | 5.41 | 9.48 | 2.28 | 18.74 | 7.86 |
| 2012 | Control | 19.49 | 2.77 | 79.12 | 4.66 | 9.76 | 2.41 | 17.17 | 7.11 |
| 2012 | Case | 19.48 | 2.88 | 79.14 | 4.69 | 9.83 | 2.52 | 17.23 | 7.22 |
| 2013 | Control | 21.53 | 3.26 | 73.53 | 5.85 | 10.95 | 3.51 | 18.44 | 9.29 |
| 2013 | Case | 21.49 | 3.30 | 73.44 | 5.88 | 11.01 | 3.52 | 18.49 | 9.30 |
| 2014 | Control | 21.08 | 2.71 | 74.24 | 5.09 | 9.18 | 2.16 | 17.21 | 8.25 |
| 2014 | Case | 21.12 | 2.73 | 74.31 | 5.11 | 9.23 | 2.17 | 17.29 | 8.30 |
| 2015 | Control | 20.33 | 2.70 | 72.86 | 5.85 | 7.66 | 1.98 | 17.11 | 7.01 |
| 2015 | Case | 20.38 | 2.70 | 72.79 | 5.86 | 7.71 | 1.98 | 17.11 | 7.00 |
| 2016 | Control | 20.79 | 2.85 | 77.04 | 5.58 | 7.47 | 2.86 | 16.18 | 7.59 |
| 2016 | Case | 20.85 | 2.87 | 77.13 | 5.51 | 7.64 | 3.00 | 16.35 | 7.77 |
| 2017 | Control | 20.88 | 2.66 | 75.99 | 4.67 | 7.27 | 2.50 | 14.95 | 7.14 |
| 2017 | Case | 20.93 | 2.74 | 75.86 | 4.65 | 7.28 | 2.53 | 14.95 | 7.20 |
| 2018 | Control | 23.05 | 3.32 | 69.97 | 8.44 | 8.24 | 3.04 | 14.55 | 6.78 |
| 2018 | Case | 23.07 | 3.37 | 69.97 | 8.65 | 8.35 | 3.06 | 14.65 | 6.79 |
| 2019 | Control | 21.31 | 3.27 | 75.85 | 5.67 | 7.05 | 2.49 | 12.99 | 6.05 |
| 2019 | Case | 21.34 | 3.37 | 75.86 | 5.71 | 7.16 | 2.65 | 13.12 | 6.15 |
| 2020 | Control | 21.06 | 3.65 | 73.69 | 8.10 | 6.59 | 3.33 | 10.46 | 4.77 |
| 2020 | Case | 21.26 | 3.75 | 73.70 | 8.24 | 6.82 | 3.69 | 10.65 | 5.01 |
| 2021 | Control | 21.15 | 2.63 | 77.01 | 4.83 | 7.54 | 2.41 | 10.83 | 4.94 |
| 2021 | Case | 21.17 | 2.64 | 77.03 | 4.90 | 7.57 | 2.43 | 10.91 | 4.97 |

**Table S2. Relative risks (95% CI) of COPD hospital admissions at the 90th, 95th, and 99th percentiles of maximum temperature and relative humidity, compared with the minimum-risk exposure, stratified by age and sex**

| Sex | Age | Temperature | | | Relative humidity | | |
| --- | --- | --- | --- | --- | --- | --- | --- |
|  |  | 90% | 95% | 99% | 90% | 95% | 99% |
| Males | 0-64 | 1.10  (1.06, 1.15) | 1.13  (1.05, 1.21) | 1.15  (0.96, 1.37) | 1.05  (1.00, 1.10) | 1.01  (0.93, 1.09) | 0.92  (0.76, 1.08) |
| Males | 65-74 | 1.11  (1.06, 1.15) | 1.16  (1.09, 1.23) | 1.29  (1.11, 1.50) | 1.07  (1.02, 1.12) | 1.09  (1.01, 1.16) | 1.12  (0.97, 1.30) |
| Males | ≥75 | 1.12  (1.08, 1.16) | 1.23  (1.17, 1.30) | 1.48  (1.30, 1.68) | 1.06  (1.01, 1.10) | 1.05  (0.99, 1.12) | 1.04  (0.90, 1.18) |
| Males | Total | 1.11  (1.09, 1.14) | 1.18  (1.14, 1.22) | 1.32  (1.21, 1.45) | 1.06  (1.03, 1.09) | 1.05  (1.01, 1.09) | 1.03  (0.95, 1.12) |
| Females | 0-64 | 1.14  (1.09, 1.19) | 1.25  (1.16, 1.33) | 1.50  (1.27, 1.77) | 1.09  (1.04, 1.14) | 1.14  (1.06, 1.22) | 1.28  (1.07, 1.53) |
| Females | 65-74 | 1.15  (1.11, 1.20) | 1.28  (1.20, 1.37) | 1.56  (1.34, 1.82) | 1.10  (1.06, 1.15) | 1.11  (1.04, 1.19) | 1.14  (0.97, 1.33) |
| Females | ≥75 | 1.14  (1.10, 1.18) | 1.25  (1.18, 1.32) | 1.52  (1.32, 1.74) | 1.06  (1.02, 1.10) | 1.05  (0.99, 1.12) | 1.02  (0.90, 1.17) |
| Females | Total | 1.15  (1.12, 1.17) | 1.26  (1.22, 1.31) | 1.54  (1.40, 1.67) | 1.08  (1.06, 1.11) | 1.09  (1.06, 1.14) | 1.12  (1.03, 1.22) |
| Total | 0-64 | 1.12  (1.09, 1.15) | 1.18  (1.13, 1.24) | 1.31  (1.16, 1.50) | 1.07  (1.03, 1.10) | 1.07  (1.01, 1.13) | 1.08  (0.96, 1.22) |
| Total | 65-74 | 1.13  (1.10, 1.16) | 1.22  (1.17, 1.27) | 1.41  (1.26, 1.57) | 1.08  (1.05, 1.12) | 1.10  (1.05, 1.15) | 1.13  (1.01, 1.26) |
| Total | ≥75 | 1.13  (1.11, 1.16) | 1.24  (1.19, 1.30) | 1.51  (1.36, 1.66) | 1.06  (1.03, 1.09) | 1.05  (1.01, 1.10) | 1.03  (0.93, 1.12) |
| Total | Total | 1.13  (1.11, 1.15) | 1.22  (1.19, 1.25) | 1.42  (1.34, 1.52) | 1.07  (1.05, 1.09) | 1.07  (1.04, 1.10) | 1.08  (1.00, 1.15) |

**Table S3. Relative risks (95% CI) of COPD hospital admissions across categories of maximum temperature and relative humidity**

| Temperature | Relative humidity | Males | Females | Total |
| --- | --- | --- | --- | --- |
| Low | Low | 1 | 1 | 1 |
| Low | Middle | 1.02 (1.00, 1.04) | 1.01 (1.00, 1.03) | 1.02 (1.00, 1.03) |
| Low | High | 1.03 (1.01, 1.05) | 1.03 (1.01, 1.04) | 1.03 (1.02, 1.04) |
| Middle | Low | 1.02 (1.00, 1.04) | 1.00 (0.98, 1.02) | 1.01 (0.99, 1.02) |
| Middle | Middle | 1.03 (1.01, 1.05) | 1.04 (1.02, 1.06) | 1.03 (1.02, 1.05) |
| Middle | High | 1.05 (1.03, 1.08) | 1.02 (1.00, 1.04) | 1.04 (1.02, 1.06) |
| High | Low | 1.08 (1.06, 1.10) | 1.05 (1.04, 1.07) | 1.06 (1.05, 1.08) |
| High | Middle | 1.08 (1.05, 1.11) | 1.08 (1.05, 1.11) | 1.08 (1.06, 1.10) |
| High | High | 1.07 (1.04, 1.11) | 1.06 (1.03, 1.10) | 1.07 (1.04, 1.09) |

Footnote: *Maximum temperature and relative humidity were categorised into three groups based on percentiles: low (<50th), medium (50th–75th), and high (≥75th).*

**Figure S1. Sensitivity analysis of exposure–response associations after additional adjustment for rainfall and wind speed**
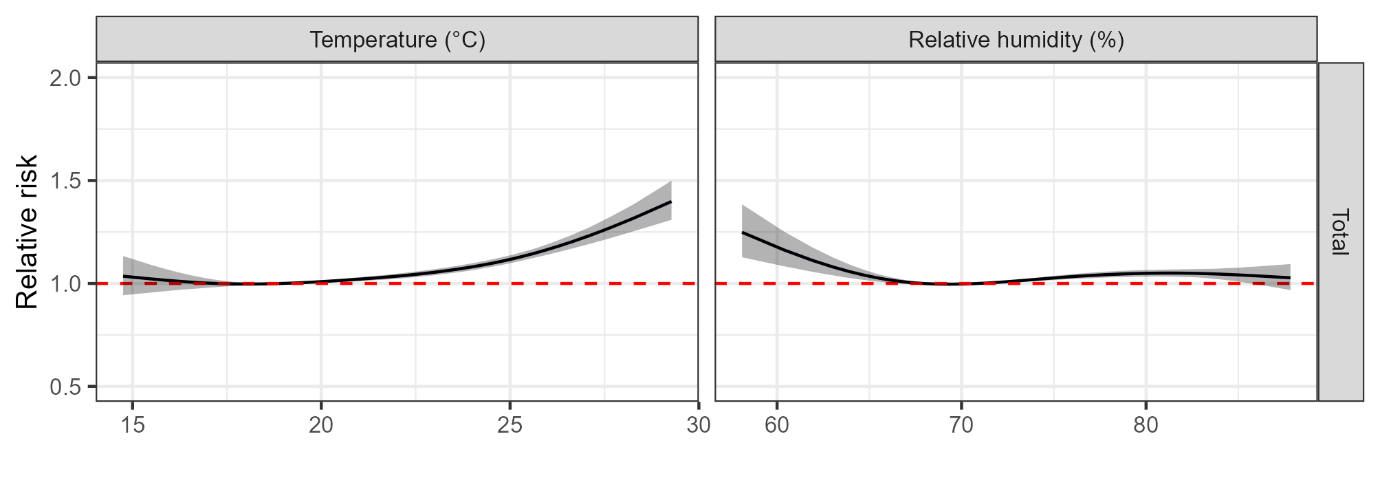

Footnote: Mean and 95% confidence intervals of the relative chronic obstructive pulmonary disease (COPD) hospital admission risk in relation with temperature and relative humidity. Results are relative to the minimum hospital admission threshold and adjusted for recurrent hospital admissions, national holidays, wind speed, rainfall and temperature or relative humidity.

**Figure S2. Sensitivity analysis of exposure–response associations using extended lag structures (0–5 days)**

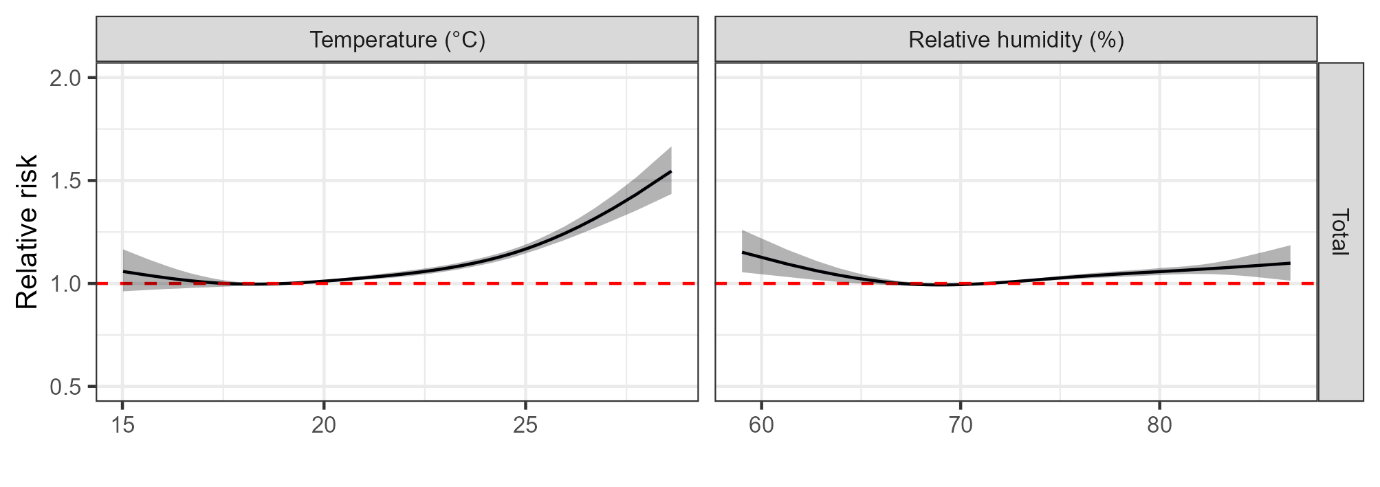

Footnote: Mean and 95% confidence intervals of the relative chronic obstructive pulmonary disease (COPD) hospital admission risk in relation with 0-5 lagged temperature and relative humidity. Results are relative to the minimum hospital admission threshold and adjusted for recurrent hospital admissions, national holidays and temperature or relative humidity.
